# Model uncertainty quantification: A post hoc calibration approach for heart disease prediction

**DOI:** 10.1101/2025.09.28.25336834

**Authors:** Peter Adebayo Odesola, Adewale Alex Adegoke, Idris Babalola

**Affiliations:** Southampton Solent University, Southampton, United Kingdom; Westminster Foundation for Democracy London, United Kingdom; Department of Health and Social Care, London, United Kingdom

**Keywords:** heart disease prediction, machine learning, probability calibration, isotonic regression, Platt scaling, uncertainty quantification, expected calibration error (ECE), Brier score, log loss, Spiegelhalter’s test, reliability diagram, post hoc calibration

## Abstract

We investigate whether post-hoc calibration improves the clinical trustworthiness of heart-disease predictions beyond conventional accuracy metrics. Using a structured clinical dataset (1,025 records; 85/15 train-test split), we benchmarked six classifiers logistic regression, SVM, k-nearest neighbors, naïve Bayes, random forest, and XGBoost on accuracy, ROC-AUC, precision, recall, and F1, and then evaluated probability quality before and after Platt (sigmoid) and isotonic calibration using Brier score, expected calibration error (ECE), log loss, Spiegelhalter’s Z-test, and reliability diagrams. Baseline discrimination was high (e.g., SVM: accuracy 92.9%, ROC-AUC 99.4%, F1 92.8%), and ensembles achieved perfect test-set scores (random forest and XGBoost: 100% across metrics), prompting calibration analysis. Isotonic calibration consistently improved probability quality for most models: random forest Brier from 0.007 to 0.002, ECE 0.051 to 0.011, log loss 0.056 to 0.012; naive Bayes Brier from 0.162 to 0.132, ECE 0.145 to 0.118, log loss 1.936 to 0.446; SVM ECE 0.086 to 0.044 and log loss 0.142 to 0.133. Platt scaling helped some models but occasionally worsened calibration (e.g., KNN ECE 0.035 to 0.081). Reliability diagrams corroborated these trends, with isotonic yielding curves closer to the 45° diagonal line, while Spiegelhalter’s test moved toward non-significance for several models’ post-calibration. Ultimately, Isotonic calibration delivered the most consistent gains in probability reliability while preserving discrimination, strengthening the interpretability and clinical actionability of model outputs.

## 1. Introduction

### 1.1. Background

Heart disease continues to be the major leading cause of death globally. It was recorded that heart disease was responsible for an estimated 19.8 million deaths in 2022 [1]. However, early and accurate prediction plays a significant role in the prevention of adverse results and reduction in healthcare costs. Machine learning (ML) models are increasingly adopted for diagnostic and prognostic tasks in cardiology due to their ability to uncover complex patterns in large clinical datasets [2].

Early ML research on heart disease cohorts primarily focused on classification accuracy, with studies routinely reporting performance above 97% using supervised classifiers [3]. These models have the capacity to learn non-linear relationships and high-dimensional interactions between contributing factors such as age, cholesterol, blood pressure, and electrocardiogram results. For example, algorithms such as Random Forest and Gradient Boosting have demonstrated superior performance to identify subtle indicators of cardiovascular abnormalities compared to traditional rule-based systems [4]. This makes them powerful techniques for risk stratification and preventive care.

However, there could be possibility that the models often provide high predictive performance, while probabilistic outputs can be poorly calibrated. That is, the confidence scores they assign do not always align with actual probabilities of disease presence [5]. In high-stakes domains such as healthcare system, well-calibrated predictions are more important to guide the appropriate treatment decisions and manage clinical risks efficiently. Miscalibrated models may lead to overconfident or underconfident decisions, ultimately compromising patient safety [6]. This has prompted a growing interest in uncertainty quantification and post hoc calibration methods, which can adjust the model’s output probabilities without retraining the original model [7]. The importance of these methods has increased in response to an increasing demand for transparent and trustworthy AI systems in clinical settings, particularly with the rise of explainable AI initiatives [8].

Furthermore, recent research has proven that visual tools such as reliability diagrams and calibration metrics such as Expected Calibration Error (ECE), Brier score, and log loss are important in evaluating how well a model is calibrated [9]. While accuracy and AUROC (Area Under the Receiver Operating Characteristic curve) remain popular metrics for model evaluation, they are insufficient for assessing how well a model estimates uncertainty. These metrics provide both quantitative and visual representations of uncertainty and prediction quality, which are vital for gaining the confidence of clinical stakeholders.

### 1.2 Motivation and Problem Statement

One of the major challenges faced by the medical health sector is the inability to detect early stages of problems related to the heart. When making decisions in the clinical sector, uncalibrated predictions may be misleading. For example, if a model predicts that a patient has a 90% chance of developing heart disease, clinicians must trust that this probability truly reflects clinical reality, otherwise this could lead to incorrect decisions and poor outcomes for the patient.

In many studies, calibration and uncertainty quantification in medical AI systems are often overlooked, leading to a gap between predictive performance and clinical trust [6]. However, this paper addresses that gap by evaluating the calibration of several popular classifiers using post hoc techniques.

### 1.3 Scope and Contributions

This study aims to evaluate and compare uncertainty estimation of heart disease prediction models. The research is guided by the following questions:

1. How do post-hoc calibration methods (Platt scaling and isotonic regression) affect the uncertainty, calibration quality, and prediction confidence of machine learning models for heart disease classification?

2. What are the baseline levels of calibration and uncertainty (ECE, Brier score, log loss, Spiegelhalter’s Z-score) for heart disease prediction before and after post-hoc calibration?

3. How does each model (e.g., Random Forest, XGBoost, SVM, KNN and Naive Bayes) perform in terms of probability calibration for heart disease before and after applying post hoc calibration?

Below, we delineate the contributions of this work in light of the research questions above. We conduct a systematic, model-agnostic evaluation of post-hoc calibration for heart-disease prediction, quantifying how Platt (sigmoid) and isotonic mapping alter probability quality without retraining the base models. Beyond headline discrimination metrics, we emphasize clinically relevant probability fidelity, calibration, sharpness, and statistical goodness-of-fit. This study makes four (4) contributions, summarized as follows:

1. A side-by-side pre/post analysis of six machine learning classifiers using reliability diagrams plus Brier, ECE, log loss, Spiegelhalter’s Z/p, and sharpness to provide complementary views of probability quality for heart disease prediction.

2. Empirical demonstration that isotonic calibration most consistently improves probability estimates, whereas Platt scaling helps some models but can worsen others.

3. Despite perfect test-set discrimination for some model, reliability diagrams reveal overconfidence pre-calibration, demonstrating why discrimination alone is insufficient for clinical use.

4. Analysis of variance in predicted probabilities shows calibration-induced smoothing and overconfidence correction, clarifying confidence reliability trade-offs relevant to clinical interpretation.

### 1.4 Related Works

#### 1.4.1 Machine Learning in Heart Disease Prediction: Calibration and Reliability Considerations

Machine learning (ML) techniques have been widely applied to predict cardiovascular disease outcomes, typically using patient risk factor data to classify the presence or risk of heart disease. For example, in heart disease prediction using supervised machine learning algorithms: Performance analysis and comparison, [10] evaluated several classifiers (KNN, decision tree, random forest, etc.) on a Kaggle heart disease dataset. They reported perfect performance with random forests achieving 100% accuracy (along with 100% sensitivity and specificity). However, their evaluation emphasized accuracy and did not include any probability calibration or uncertainty quantification. Similarly, [11] evaluation of Heart Disease Prediction Using Machine Learning Methods with Elastic Net Feature Selection compared logistic regression (LR), KNN, SVM, random forest (RF), AdaBoost, artificial neural network (ANN), and multilayer perceptron on the Kaggle dataset used in this study. They found RF to attain ∼99% accuracy and AdaBoost ∼94% on the full feature set and observed SVM performing best after SMOTE class-balancing and feature selection. Like [10], this study focused on accuracy improvements and other discrimination metrics, with no model calibration applied.

Another work by [12], they also utilized the Kaggle dataset we explored. They evaluated a wide range of classifiers including RF, decision tree (DT), gradient boosting (GBM), KNN, AdaBoost, LR, ANN, QDA, LDA, SVM and reported extremely high accuracy for ensemble methods. In fact, their RF model reached 100% training accuracy (and ∼99% under cross-validation). Despite reporting precision, recall, F1-score, and ROC-AUC for each model, this work too did not report any calibration metrics or uncertainty estimates; the focus remained on discrimination performance.

Beyond the popular Kaggle/UCI datasets, researchers have explored ML on other heart disease cohorts. For instance, [13] in A Machine Learning Model for Detection of Coronary Artery Disease s applied ML to the Z-Alizadeh Sani dataset (303 patients from Tehran’s Rajaei cardiovascular center). They employed six algorithms (DT, deep neural network, LR, RF, SVM, and XGBoost) to predict coronary artery disease (CAD). After Pearson-correlation feature selection, the best results were achieved by SVM and LR, each attaining 95.45% accuracy with 95.91% sensitivity, 91.66% specificity, F1≈0.969, and AUROC ≈0.98. Notably, although this study achieved excellent discrimination, it did not incorporate any post-hoc probability calibration or uncertainty analysis, the evaluation centered on accuracy and ROC curves alone.

[14] took a different approach by leveraging larger, real-world data. In an interpretable LightGBM model for predicting coronary heart disease: Enhancing clinical decision-making with machine learning, they trained a LightGBM model on a U.S. CDC survey dataset (BRFSS 2015) and validated on two external cohorts (the Framingham Heart Study and the Z-Alizadeh Sani data). The LightGBM achieved about 90.6% accuracy (AUROC ∼81.1%) on the BRFSS training set, with slightly lower performance on Framingham (85% accuracy, ∼67% AUROC) and Z-Alizadeh (80% accuracy). While Deng et al. prioritized model interpretability (using SHAP values) and reported standard metrics like accuracy, precision, recall, and AUROC, they did not report any calibration-specific metrics (e.g. no ECE, Brier score, or reliability diagrams), nor did they apply Platt scaling or isotonic regression in their pipeline. Several recent studies have pushed accuracy to very high levels by combining datasets or using advanced ensembles, yet still largely ignore calibration. [15] proposed a hybrid approach for predicting heart disease using machine learning and an explainable AI method, where they combined a private hospital dataset with a public one and used feature selection plus ensemble methods. Their best model (an XGBoost classifier on a selected feature subset SF-2) achieved 97.57% accuracy with 96.61% sensitivity, 90.48% specificity, 95.00% precision, F1=92.68%, and 98% AUROC. Despite this impressive performance, no probability calibration was mentioned; the study’s contributions focused on maximizing accuracy and explaining feature impacts (via SHAP) rather than assessing prediction uncertainty.

Using a clinical and biometric dataset (n=571) with a man-in-the-loop paradigm for assessing coronary artery disease, [16] compared standard ML classifiers; best accuracy reached ≈83% with expert input, but the work emphasized explainability over probabilistic calibration. To address the need for diverse and comprehensive research, we conducted a lightweight systematic review and surveyed a range of peer reviewed studies on ML for heart disease prediction in the last 5–10 years with focus on a minimum of 5,000 cohort patients built into the experimental setup. Table 1 summarizes key studies, including their data sources, ML approaches, and whether model calibration was evaluated (and how).

**Table 1.** Recent ML-based heart disease prediction studies (2017-2025) - Summary of data, methods, and calibration evaluation. (Calibration metrics: HL = Hosmer–Lemeshow test; ECE = Expected Calibration Error; O/E = observed-to-expected ratio; Brier = Brier score.)

| Year [Ref] | Data (Population / Dataset) | ML Approach & Key Results | Calibration (Evaluation & Metrics) |
| --- | --- | --- | --- |
| 2025 [17] | Japanese Suita cohort (n=7,260; ~15-year follow-up; ages 30-84). | Risk models (LR, RF, SVM, XGB, LGBM) for 10 year CHD; RF best (AUC ~0.73); SHAP identified key factors. | Yes - Calibration curves and O/E ratios; RF ~1:1 calibration. |
| 2025 [18] | NHANES (USA; ~37,000). | PSO ANN - particle swarm optimized neural net; ~97% accuracy; surpassed LR (~95.8%); feature selection + SMOTE. | No - Calibration not reported. |
| 2024 [19] | Simulated big dataset + UCI. | AttGRU HMSI deep model; ~95.4% accuracy; emphasis on big data processing and feature selection. | No - Calibration not reported. |
| 2023 [20] | UK Biobank (n≈473,000; 10 year follow up). | AutoPrognosis AutoML; AUC ≈0.76; 10 key predictors discovered. | Yes - Brier ~0.057 (good calibration). |
| 2023 [21] | China EHR (Ningbo; n=215,744; 5 year follow up). | XGBoost vs Cox; C index 0.792 vs 0.781. | Yes - HL $\chi^2$ ≈0.6, p=0.75 in men; non significant HL (good calibration). |
| 2023 [22] | Stanford ECG datasets; external validation at 2 hospitals. | SEER CNN using resting ECG; 5 yr CV mortality AUC ~0.80 - 0.83; ASCVD AUC ~0.67; reclassified ~16% low risk to higher risk with true events. | No - Calibration not reported. |
| 2022 [23] | China hypertension cohort (n=143,043). | Ensemble (avg RF/XGB/DNN); AUC 0.760 vs LR 0.737. | No - Calibration not reported. |
| 2021 [24] | Korea NHIS (n≈223k) + external cohorts. | ML vs risk scores for 5 yr CVD; simple NN improved C stat (0.751 vs 0.741). | Yes - HL $\chi^2$ baseline 171 vs 15-86 for ML (p>0.05). Brier ~0.031 - 0.032 (good calibration). |
| 2021 [25] | NCDR Chest Pain MI registry (USA; n=755,402; derivation 564k; validation 190k). | In hospital mortality after MI; ensemble/XGBoost/NN vs logistic; similar AUC (~0.89). | Yes - Calibration slope ~1.0 in validation; Brier components & recalibration tables reported. |
| 2021 [26] | Faisalabad Institute + Framingham + South African Hearth dataset & UCI (Cleveland n=303). | Feature importance with 10 ML algorithms; XAI focus. | No - Calibration not reported. |
| 2020 [27] | Eastern China high risk screening (n=25,231; 3 year follow up). | Random Forest; AUC ≈0.787 vs risk charts ≈0.714. | Yes - HL $\chi^2$=10.31, p=0.24 (good calibration). |
| 2019 [28] | UK Biobank subset (n=423,604; 5 year follow-up). | AutoPrognosis ensemble; AUC ≈0.774 vs Framingham ≈0.724; +368 cases identified. | Yes - Pipeline includes calibration (e.g., Platt scaling [sigmoid]); good agreement of predicted vs observed risk. |
| 2017<br>[29] | UK CPRD primary care (n=378,256; 10 year follow up; 24,970 events). | Classic ML vs ACC/AHA score; NN best (AUC ≈0.764) vs 0.728; improved identification. | No - Calibration not reported. |

Each study is cited with its year and reference number (e.g., 2025 [17] means the study was published in 2025 and is reference [17] in the reference list).

### 1.4.2 Gaps in Research

Despite abundant work on ML-based heart disease prediction, there are clear gaps in the literature regarding probability calibration and uncertainty quantification. First, most studies prioritize discriminative performance (accuracy, F1, AUROC, etc.) and devote little or no attention to how well the predicted probabilities reflect true risk. As shown above, prior works seldom report calibration metrics like ECE or Brier score, nor do they plot reliability diagrams. For example, none of the 10+ studies reviewed applied calibration methods such as Platt scaling or isotonic regression to their classifiers, except for only one study [28]. This indicates a lack of focus on calibration quality, an important aspect if these models are to be used in clinical decision-making where calibrated risk predictions are crucial.

Second, there is a lack of unified evaluation across multiple models and calibration techniques. Prior research typically evaluates a set of ML models on a dataset (as in comparative studies) but stops at reporting raw performance metrics. No study to date has systematically taken multiple classification models for heart disease and evaluated them before and after post-hoc calibration. This means it remains unclear how different algorithms (e.g. an SVM vs. a random forest) compare in terms of probability calibration (not just classification accuracy), and whether simple calibration methods can significantly improve their reliability. Furthermore, the interplay between model uncertainty (e.g. variance in predictions) and calibration has not been explored in this domain. Third, most heart disease prediction papers do not report uncertainty metrics or advanced calibration statistics. Metrics such as the Brier score (which combines calibration and refinement), the ECE (Expected Calibration Error), or even more domain-specific checks like Spiegelhalter’s Z-test for calibration, are virtually absent from prior studies. Sharpness (the concentration of predictive distributions) and other uncertainty measures are also not discussed. This leaves a research gap in understanding how confident we can be in these model predictions and where they might be over or under-confident. For instance, none of the reviewed studies provide reliability diagrams to visually inspect calibration; as a result, a model claiming 95% accuracy might still make poorly calibrated predictions (overestimating or underestimating risk).

To the best of our knowledge, no prior work has offered a comprehensive evaluation of pre and post-calibration metrics across multiple models on the specific Kaggle heart disease dataset (1,025 records) used in this study. While several papers have used this or similar data for model comparison, none have examined calibration changes (ECE, log-loss, Brier, sharpness, Spiegelhalter’s Z-test, calibration curves) resulting from post-hoc calibration methods (Platt scaling, isotonic regression). In short, existing studies have left a critical question unanswered: if we calibrate our heart disease prediction models, do their confidence estimates become more trustworthy, and how does this vary by model? Addressing this gap is the focus of our work. We provide a thorough assessment of multiple classifiers before and after calibration, using a suite of calibration and uncertainty metrics not previously applied in this context, thereby advancing the evaluation criteria for heart disease ML models beyond conventional accuracy-based measures.

## 2 Materials and Methods

### 2.1 Research Methodology Overview

This study employs a structured machine learning workflow to predict heart disease risk based on clinical and demographic variables. As outlined in Figure 1, the process begins with the heart disease dataset, followed by data preprocessing, model selection and training, performance evaluation, and post-hoc calibration. Two calibration techniques (i.e Platt Scaling and Isotonic Regression) are applied to refine probabilistic outputs, with effectiveness assessed.

**Figure 1.**
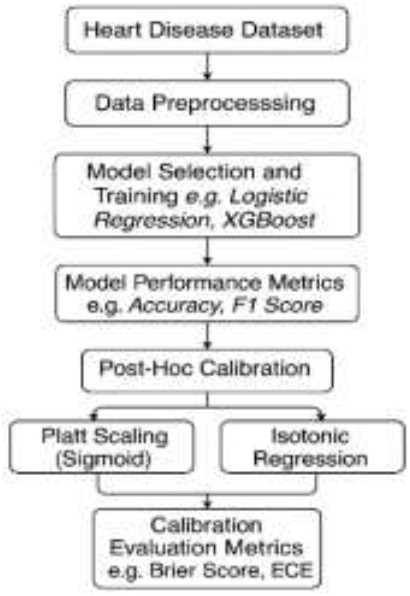
Workflow Diagram for Heart Disease Prediction and Calibration Pipeline

### 2.2 Description of Dataset

The Heart Disease dataset used in this study was sourced from Kaggle. It was originally sourced by merging data from four medical centers Cleveland, Hungary, Switzerland and VA Long Beach bringing the sample size to 1,025 records including 713 males (69.6%) and 312 females (30.4%), ages ranging between 29 - 77 years (median age ∼56). The dataset contains 14 variables encompassing demographic, clinical and diagnostic test features. Descriptions of the dataset are outlined in Table 2.

**Table 2.** Data description for heart disease.

| Feature | Description | Data Type | Values / Range |
| --- | --- | --- | --- |
| Age(Years) | Age of the patient | Integer | 29–77 |
| sex | Sex (1 = male, 0 = female) | Categorical | 0, 1 |
| cp | Chest pain type | Categorical | 1: typical angina, 2: atypical angina, 3: non-anginal pain, 4: asymptomatic |
| trestbps(mmHg) | Resting blood pressure (on admission to the hospital) | Integer | 94–200 |
| chol(mmol/L) | Serum cholesterol | Integer | 126–564 |
| Fbs (mmol/L) | Fasting blood sugar > 120 mg/dl (1 = true, 0 = false) | Categorical | 0, 1 |
| restecg | Resting electrocardiographic results | Categorical | 0: normal, 1: ST-T abnormality, 2: left ventricular hypertrophy |
| thalach | Maximum heart rate achieved | Integer | 71–202 |
| exang | Exercise induced angina (1 = yes, 0 = no) | Categorical | 0, 1 |
| oldpeak | ST depression induced by exercise relative to rest | Real | 0.0–6.2 |
| slope | Slope of the peak exercise ST segment | Categorical | 1: upsloping, 2: flat, 3: downsloping |
| ca | Number of major vessels (0–3) colored by fluoroscopy | Integer | 0–3 |
| thal | Thalassemia test result | Categorical | 3: normal, 6: fixed defect, 7: reversible defect |
| num | Presence of heart disease (target: 0 = no, 1-4 = disease) | Categorical | 0, 1, 2, 3, 4 |

The dataset was inspected for missing values and none was identified. The outcome variable (Heart Disease) was approximately balanced, with 51.3% of records labeled Disease and 48.7% labeled No Disease as shown in Figure 2.

**Figure 2.**
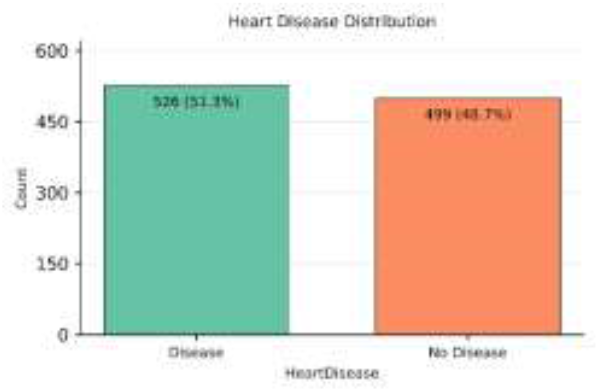
Heart disease distribution

### 2.3 Data Preprocessing

In this study, the dataset was separated into 13 predictors (i.e patient risk factors) and the 1 outcome feature (i.e the presence or risk of heart disease). Predictors were further divided into two groups: *numerical features* (e.g Age, RestingBP, Cholesterol) and *categorical features* (e.g ChestPainType, RestingECG, Thalassemia, Sex). We scale numerical features using a RobustScaler approach, which centres values around the median and spreads them according to the interquartile range. This method was selected due to it being less sensitive to outliers and skewness [30]. For categorical features, a One-Hot Encoding approach was applied, converting each category into binary (0/1) variables. This ensured that all categories were represented in a machine-readable format.

### 2.4 Model Selection

In this work, we benchmark six models (spanning linear, non-linear and ensemble model architecture) to classify patients based on the presence or absence of heart disease. Models selected include Logistic Regression (LR), Support Vector Machines (SVM), Random Forest (RF), Extreme Gradient Boosting (XGBoost), K-Nearest Neighbors (KNN), and Naive Bayes (NB). The data was split into training (85%) and testing (15%) sets. We trained each model on the preprocessed training data and evaluated on the held-out test data.

#### Logistic Regression (LR)

Logistic Regression is a supervised machine learning model well-suited for binary classification, such as determining the presence or absence of heart disease. LR calculates the probability of a class (e.g., disease or no disease) by applying a sigmoid function to a weighted sum of predictor variables. Its strengths include simplicity, efficiency, and the ability to interpret coefficients as odds ratios, which is valuable in clinical settings for understanding feature importance and risk factors. Logistic Regression has a proven track record in medical research for risk stratification and is easily calibrated for probability estimation [31].

#### Support Vector Machines (SVM)

Support Vector Machines are powerful, supervised classification models that work by finding the optimal hyperplane that separates classes in the feature space. SVMs excel at handling high-dimensional data and can model nonlinear relationships through kernel tricks, making them highly effective for complex medical datasets. Their ability to maximize the margin between classes reduces the likelihood of misclassification, which is especially useful when distinguishing subtle differences between patients with and without heart disease. SVMs are known for their robustness in real-world clinical prediction tasks [32]

#### Random Forest (RF)

Random Forest is an ensemble algorithm that builds multiple decision trees during training and aggregates their outputs via majority voting for classification. It is especially effective at capturing nonlinear relationships and interactions among risk factors in heart disease prediction. The ensemble nature of RF mitigates overfitting and variance, providing more reliable and stable predictions on diverse patient populations. Its embedded feature importance scores help clinicians identify key predictors of heart disease, further supporting its use in healthcare analytics [33]

#### Extreme Gradient Boosting (XGBoost)

XGBoost is a gradient boosting framework that creates a series of weak learners (usually decision trees) and optimizes them sequentially. It is renowned for combining high predictive accuracy with speed and efficiency, making it a top performer in medical classification challenges. XGBoost handles missing data gracefully and is robust to outliers, both common in clinical datasets. Its sophisticated regularization techniques reduce overfitting, and its model interpretability tools are advantageous for validating results in heart disease risk prediction [34].

#### K-Nearest Neighbors (KNN)

K-Nearest Neighbors is a non-parametric classification method that predicts the class of a sample based on the majority class among its k closest neighbors in feature space. KNN is intuitive, easy to implement, and doesn’t make assumptions about data distribution, making it suitable for heterogeneous clinical datasets. KNN is effective at leveraging local patterns, which can be helpful for identifying at-risk heart disease patients based on similarity to previously observed cases. However, it can be sensitive to feature scaling and less efficient with very large datasets [35].

#### Naive Bayes (NB)

Naive Bayes is a probabilistic classification algorithm that applies Bayes’ theorem, assuming feature independence. Its simplicity and computational efficiency make it attractive for medical tasks with many categorical variables. Despite its “naive” independence assumption, NB often performs surprisingly well for heart disease prediction because it can handle missing values, is robust with noisy data, and quickly estimates posterior probabilities. This makes it valuable for real-time risk assessment and decision support in clinical environments [36].

### 2.5 Model Performance Metric

We evaluated classification performance using Accuracy, ROC-AUC, Precision, Recall, and F1-score. Let TP, FP, TN, and FN denote true positives, false positives, true negatives, and false negatives, respectively.

#### Accuracy

Defined as 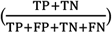, accuracy reflects the share of correctly classified cases in the test set. In clinical screening contexts where disease prevalence may be low accuracy depends on the decision threshold and can mask deficiencies under class imbalance, yielding seemingly strong performance while missing many positive cases [37].

#### ROC-AUC

The receiver-operating-characteristic area summarizes discrimination across all thresholds; it equals the probability that a randomly selected positive receives a higher score than a randomly selected negative and ranges from 0.5 (no discrimination) to 1.0 (perfect). ROC-AUC is broadly used in clinical prediction for its threshold-agnostic view of separability, though it does not reflect calibration or the clinical costs of specific error types [38].

#### Precision

Given by 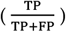, quantifies how reliable positive alerts are among patients flagged as having heart disease, the fraction truly positive. As thresholds are lowered to capture more cases, precision typically decreases, illustrating the trade-off clinicians face between false alarms and case finding [39].

#### Recall

Defined as 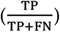, measures the proportion of truly diseased patients the model detects (sensitivity). Raising recall generally requires a lower threshold, which increases false positives and reduces precision; selecting an operating point should therefore reflect clinical consequences and disease prevalence [40].

#### F1-score

The harmonic mean 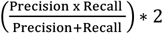,provides a single summary when both missed cases and false alarms matter. F1 is commonly reported in imbalanced biomedical tasks, though its interpretation should be complemented by other metrics given known limitations under skewed prevalence [41].

These metrics establish a consistent baseline for cross-model comparison and inform our subsequent calibration and uncertainty quantification analysis.

### 2.6 Post-Hoc Calibration Techniques

Post-hoc calibration refers to techniques applied after model training that map raw scores to probabilities without changing the underlying classifier. In clinical settings where decisions hinge on risk estimates, these procedures use a held-out calibration set to fit a simple, typically monotonic mapping so that predicted probabilities better match observed event rates [9][42] [43]. In clinical text or imaging pipelines for heart-disease prediction, this is attractive: one can retain the trained model and its operating characteristics, then calibrate its outputs to yield probabilities that are more trustworthy for downstream decision thresholds, alerts, or shared decision-making [42][43]. For this study, we applied two post hoc calibration methods, Platt scaling (sigmoid calibration) and isotonic regression to adjust model outputs (such as prediction scores) into well-calibrated probabilities [7].

(i) Platt scaling works by fitting a smooth S-shaped (sigmoid) curve to the model’s scores using a separate validation set, so that predicted probabilities better match actual outcomes. This method is simple and efficient but assumes that the relationship between scores and probabilities follows a logistic pattern [9][44].

(ii) Isotonic regression is a more flexible, non-parametric method that does not assume any specific shape. Instead, it fits a step-like monotonic curve that can adapt to complex patterns in the data [45]. While this flexibility can better capture irregular relationships, it can also lead to overfitting if the validation dataset is small.

In practice, Platt scaling is most useful when a sigmoid relationship is expected, while isotonic regression is preferred when the calibration pattern is unknown or more complex [9]. Using both methods together provides a robust calibration toolbox, ensuring reliable probability estimates across different models. This is particularly important in clinical applications such as heart disease prediction, where well-calibrated risk probabilities are essential for trustworthy model outputs and informed medical decision-making.

### 2.7 Model Uncertainty Quantification and Calibration Evaluation Metrics

In this study, we measure the uncertainty of the models using these key calibration evaluation metrics: Brier Score, Expected Calibration Error (ECE), Log Loss, Spiegelhalter’s Z-score & p-value, Sharpness and reliability diagram. A combination of these metrics provides a holistic understanding of the effectiveness of each model in relation to quantifying model uncertainty.

#### Brier Score

The Brier Score measures the mean squared difference between predicted probabilities and the actual binary outcomes. Unlike accuracy which reduces predictions to “yes/no” and ignores the uncertainty behind probability values the Brier Score penalizes poorly calibrated or overly confident predictions. This makes it more informative for model uncertainty quantification, especially in clinical settings were knowing the probability of heart disease (and not just a binary label) aids risk discussions and decision-making. Lower Brier Scores indicate better calibrated and more reliable probability forecasts, a key aspect of clinical utility [46].

#### Expected Calibration Error (ECE)

ECE summarizes how closely a model’s predicted probabilities match the observed frequencies of outcomes. It divides predictions into probability bins and measures mismatch between average predicted probability and actual outcome rate in each bin. In heart disease prediction, ECE helps verify if model confidence reflects real-world risks, ensuring patients with a predicted 70% heart disease risk, for example, actually face that risk. Lower ECE values indicate better calibrated models crucial for trusted clinical decision support [5].

#### Log Loss

Log Loss (or cross-entropy loss) evaluates the uncertainty of probabilistic outputs by heavily penalizing confident but incorrect predictions. Log Loss is sensitive to how far predicted probabilities diverge from the actual class, providing a continuous measure of model reliability. For heart disease prediction, low Log Loss means the model rarely makes wildly overconfident errors, promoting safer, uncertainty-aware clinical interpretation [47].

#### Spiegelhalter’s Z-score & p-value

Spiegelhalter’s Z-score tests overall calibration by comparing predicted probabilities to actual outcomes, normalized by their variance. A non-significant p-value suggests the model is well-calibrated; otherwise, the probabilistic forecasts may be systematically over- or under-confident. This calibration test is especially important in health applications, assuring clinicians that model probabilities are statistically valid reflections of true outcome chances [48].

#### Sharpness (variance of predicted probabilities)

Sharpness measures the spread or concentration of predicted probabilities, independent of whether they’re correct. High sharpness means the model often predicts risks near 0 or 1, indicating confident, decisive forecasts. For heart disease prediction, greater sharpness is desirable only if paired with good calibration confident predictions should be correct. Thus, sharpness reveals how much intrinsic uncertainty the model expresses, helping physicians judge whether predictions are actionable or too vague for clinical use [49].

#### Reliability Diagram (calibration plot)

A reliability diagram visualizes how predicted probabilities align with observed event rates by plotting, across confidence bins, the empirical outcome frequency against the mean predicted probability. A perfectly calibrated model traces the 45° diagonal line; systematic deviations reveal over or under-confidence [9]. Reliability diagrams are standard in both forecast verification and machine-learning calibration, and they are visual check of probability accuracy while leaving discrimination unaffected. Practical caveats include sensitivity to binning and sample size, and the fact that the plot alone does not convey how many samples fall in each bin often addressed by adding a companion confidence histogram.[5][49]

### 2.8 Experimental Setup

Table 3 summarizes the experiment setup, evaluation choices, and preprocessing decisions.

**Table 3.** Pipeline decisions for Baseline Classification Performance & Calibration.

| Component | Description |
| --- | --- |
| Test Split | 15% of dataset (~153 instances), stratified by target class |
| Cross-Validation | 5-fold StratifiedKFold with shuffling |
| Scaling | RobustScaler for numeric variables |
| Encoding | OneHotEncoder for nominal categorical fields |
| Models | Logistic Regression, SVM, Random Forest, XGBoost, KNN, Naive Bayes |
| Development Environment | Google Colab |
| Python libraries | Sklearn, matplotlib, scipy, numpy, pandas, seaborn |
| Model Evaluation Metrics | Accuracy, ROC-AUC, Precision, Recall, and F1 Score |
| Uncertainty Quantification Metrics | Brier Score, Expected Calibration Error (ECE), Log Loss, Spiegelhalter’s Z-score & p-value, Sharpness, Reliability diagram |
| Train/test split ratio | 85% training: 15% testing |

## 3. Results

### 31 Baseline Model Performance (Before Calibration)

To establish a performance benchmark for probability calibration, six (6) classification models were trained and evaluated. Table 4 summarizes evaluation metrics including accuracy, ROC AUC, precision, recall, F1 score, and 5-fold cross-validated accuracy (mean ± standard deviation) on the test set.

**Table 4.** Performance metrics of baseline classification models (before calibration).

| Model | Accuracy (%) | ROC AUC (%) | Precision (%) | Recall (%) | F1 Score (%) | CV Accuracy Mean (%) | CV Std |
| --- | --- | --- | --- | --- | --- | --- | --- |
| LR | 88.3 | 96.6 | 86.7 | 91.1 | 88.9 | 86.1 | 2.9 |
| SVM | 92.9 | 99.4 | 95.9 | 89.9 | 92.8 | 92.0 | 3.0 |
| RF | 100 | 100 | 100 | 100 | 100 | 98.7 | 0.9 |
| XGB | 100 | 100 | 100 | 100 | 100 | 98.2 | 1.1 |
| KNN | 88.3 | 97.3 | 87.7 | 89.9 | 88.7 | 85.0 | 4.1 |
| NB | 81.8 | 87.6 | 81.5 | 83.5 | 82.5 | 82.0 | 2.4 |

The 6 classifiers all achieved competitive results, with ensemble-based models (Random Forest and XGBoost) attaining perfect scores across all reported metrics on the held-out test set. SVM also performed robustly, yielding an accuracy of 92.9%, ROC AUC of 99.4%, and F1 score of 92.8%. Logistic Regression and KNN followed closely, whereas Naive Bayes, while comparatively lower in performance (accuracy = 81.8%), remained competitive with an F1 score of 82.5%. However, the perfect test-set performance by the ensemble models may suggest overfitting, especially given their slightly lower cross-validation accuracy (Random Forest: 98.7%; XGBoost: 98.2%), warranting further scrutiny via probabilistic calibration metrics and establishing a foundation for subsequent analysis of uncertainty quantification to determine whether high predicted probabilities genuinely reflect empirical frequencies.

### 32 Calibration and Uncertainty Before & After Post-Hoc Adjustment

We plot reliability diagrams and partitioned predicted probabilities into five equal-width bins. Given a test set of 153 instances (15% of the 1,025-record dataset), this produces approximately 30 samples per bin, striking a balance between calibration resolution and statistical stability. This aligns with prior calibration studies which recommend ≥10 bins depending on dataset size and caution against excessive binning for modestly sized datasets to maintain interpretability and reduce sampling noise [50].

As shown in Figure 3 (Before Calibration), XGBoost and Logistic Regression are close to the diagonal line, suggesting good intrinsic calibration. Naive Bayes and KNN showed substantial deviations from perfect calibration, with Naive Bayes underestimating outcome at intermediate predicted probability levels confirming the well-known tendency of these models to produce poorly calibrated outputs [7][51] and KNN displaying slightly erratic behavior, indicative of high-variance estimates [9]. Ensemble models, despite perfect accuracy, were overconfident in high-probability regions especially Random Forest where the predicted values exceeded observed outcomes. In addition, we observed that the XGBoost model’s pre-calibration reliability diagram was already close to the diagonal, indicating well-calibrated initial probabilities.

**Figure 3.**
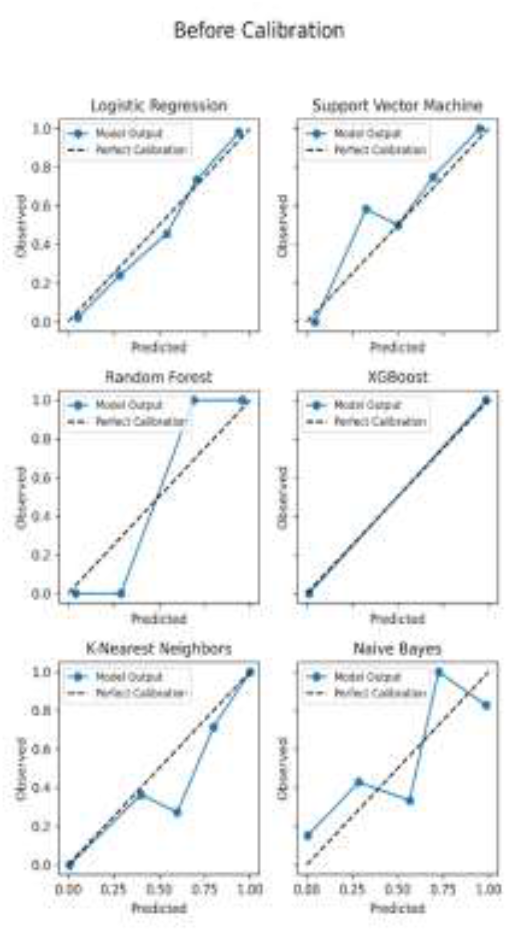
Reliability diagrams before calibra

Application of sigmoid calibration improved the calibration of Logistic Regression and SVM bringing their reliability curves closer to the diagonal (Figure 4). However, sigmoid calibration failed to correct significant non-linear calibration errors in Random Forest and KNN, likely due to its monotonic, logistic-form constraint [52][53]. Naive Bayes also showed improvement, though some mid-range under confidence persisted. We noted that the reliability curve became more erratic for XGBoost which distorted the well-calibrated probabilities in Figure 3 rather than improving them suggesting additional calibration was not beneficial.

**Figure 4.**
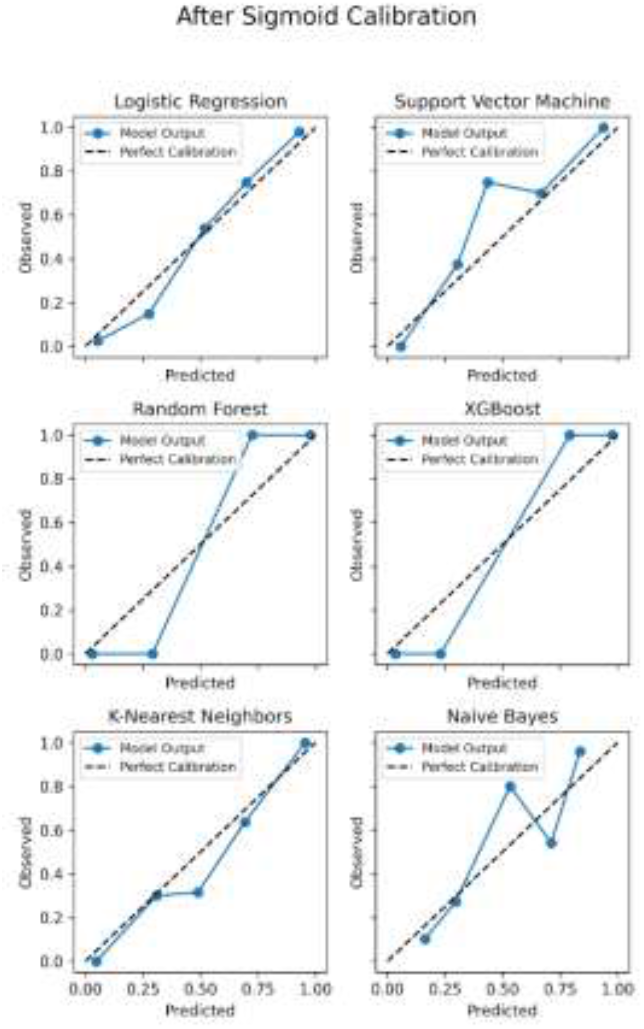
Reliability diagrams after sigmoid calibration

In contrast, isotonic calibration produced more flexible corrections (Figure 4), significantly aligning the predictions of SVM and Naive Bayes with observed event frequencies. Whereas KNN again remained unstable across bins. These findings suggest that while sigmoid calibration is suitable for models with nearly linear miscalibration, isotonic regression better handles complex, non-monotonic distortions in probabilistic estimates [54][55].

**Figure 5.**
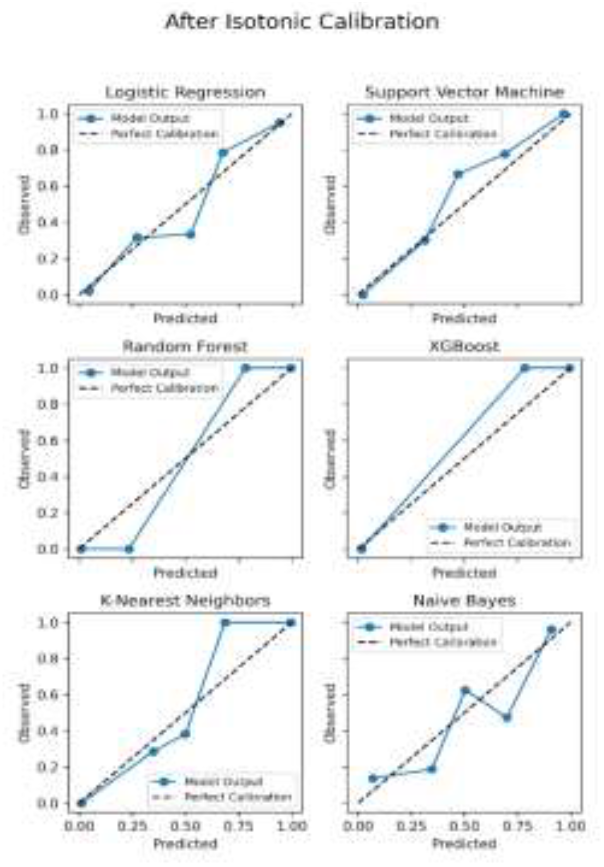
Reliability diagrams after isotonic calibration

To complement the visual insights from reliability diagrams, we conducted a quantitative evaluation using six (6) calibration metrics: *Brier Score, Expected Calibration Error (ECE), Log Loss, Sharpness (Variance), Spiegelhalter’s Z-test and Spiegelhalter’s P-value* shown in Table 5, to quantify calibration quality and predictive uncertainty before and after applying sigmoid and isotonic calibration.

**Table 5.**
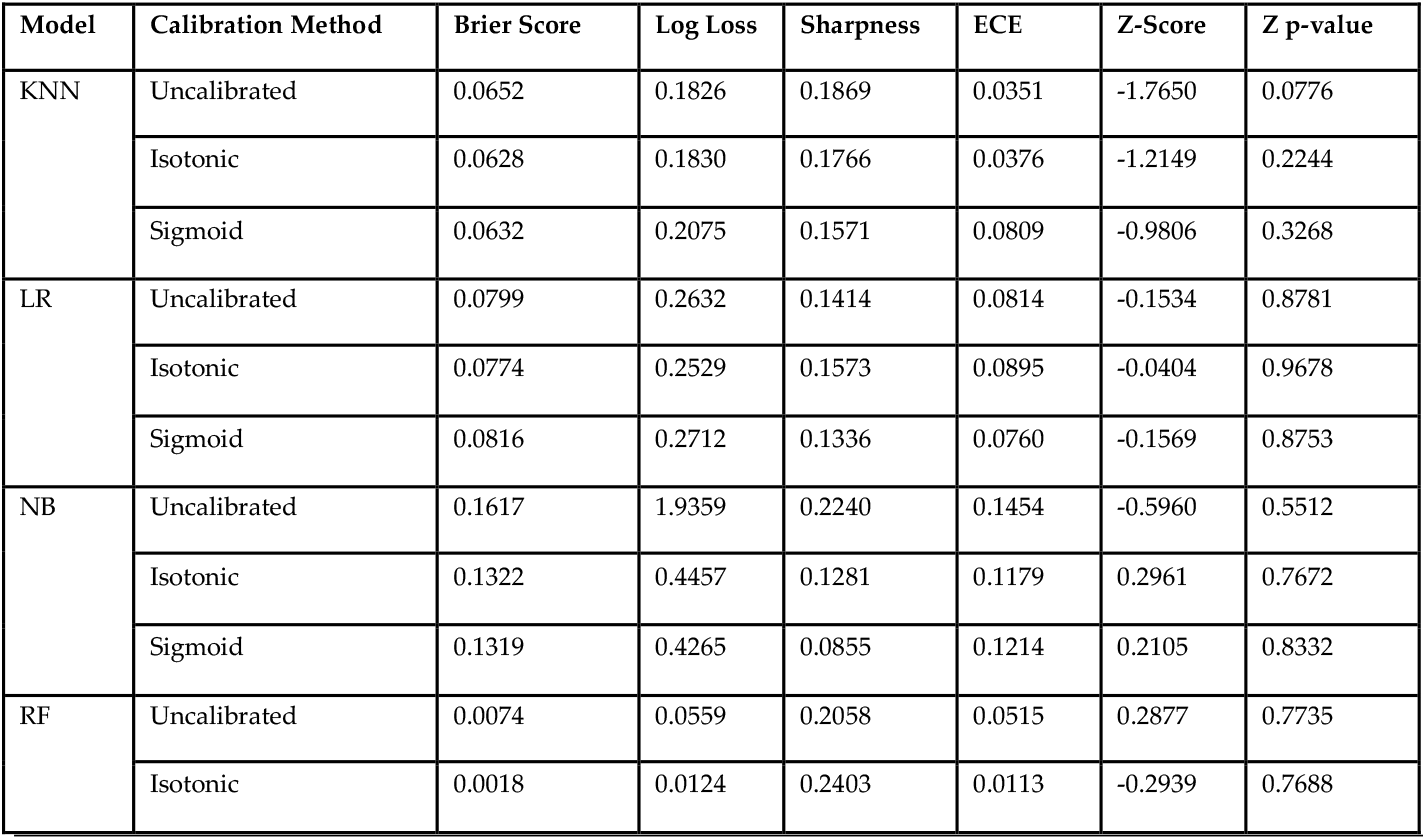
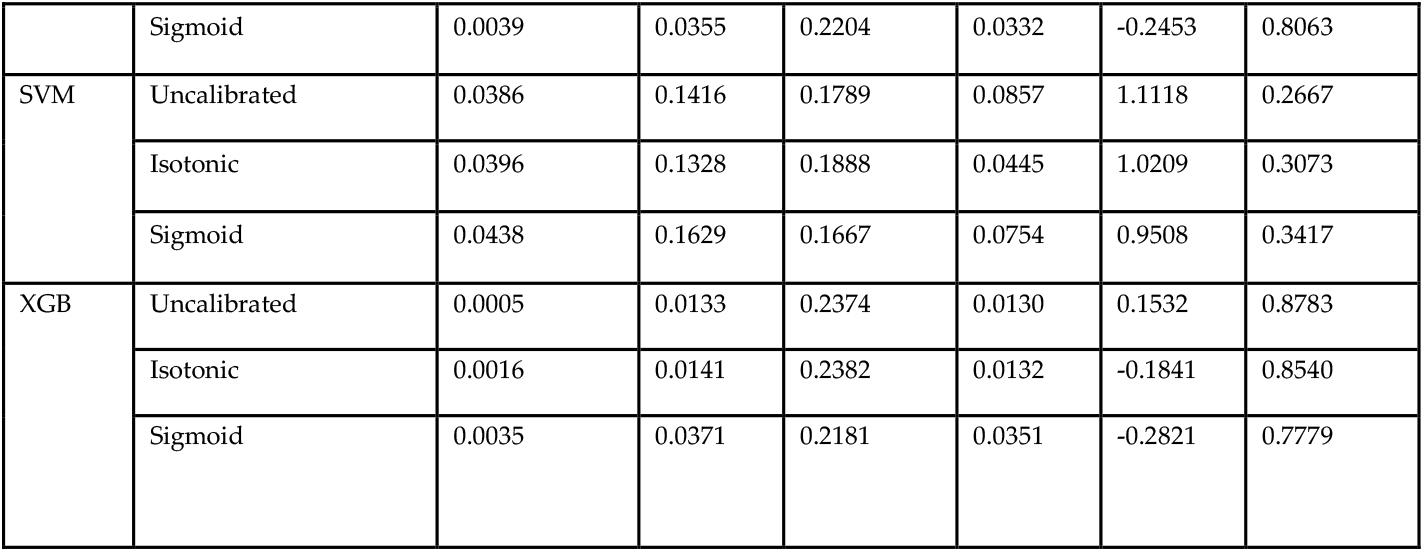
Calibration Metrics by Model & Calibration Method.

| Model | Calibration Method | Brier Score | Log Loss | Sharpness | ECE | Z-Score | Z p-value |
| --- | --- | --- | --- | --- | --- | --- | --- |
| KNN | Uncalibrated | 0.0652 | 0.1826 | 0.1869 | 0.0351 | -1.7650 | 0.0776 |
|  | Isotonic | 0.0628 | 0.1830 | 0.1766 | 0.0376 | -1.2149 | 0.2244 |
|  | Sigmoid | 0.0632 | 0.2075 | 0.1571 | 0.0809 | -0.9806 | 0.3268 |
| LR | Uncalibrated | 0.0799 | 0.2632 | 0.1414 | 0.0814 | -0.1534 | 0.8781 |
|  | Isotonic | 0.0774 | 0.2529 | 0.1573 | 0.0895 | -0.0404 | 0.9678 |
|  | Sigmoid | 0.0816 | 0.2712 | 0.1336 | 0.0760 | -0.1569 | 0.8753 |
| NB | Uncalibrated | 0.1617 | 1.9359 | 0.2240 | 0.1454 | -0.5960 | 0.5512 |
|  | Isotonic | 0.1322 | 0.4457 | 0.1281 | 0.1179 | 0.2961 | 0.7672 |
|  | Sigmoid | 0.1319 | 0.4265 | 0.0855 | 0.1214 | 0.2105 | 0.8332 |
| RF | Uncalibrated | 0.0074 | 0.0559 | 0.2058 | 0.0515 | 0.2877 | 0.7735 |
|  | Isotonic | 0.0018 | 0.0124 | 0.2403 | 0.0113 | -0.2939 | 0.7688 |
|  | Sigmoid | 0.0039 | 0.0355 | 0.2204 | 0.0332 | -0.2453 | 0.8063 |
| SVM | Uncalibrated | 0.0386 | 0.1416 | 0.1789 | 0.0857 | 1.1118 | 0.2667 |
|  | Isotonic | 0.0396 | 0.1328 | 0.1888 | 0.0445 | 1.0209 | 0.3073 |
|  | Sigmoid | 0.0438 | 0.1629 | 0.1667 | 0.0754 | 0.9508 | 0.3417 |
| XGB | Uncalibrated | 0.0005 | 0.0133 | 0.2374 | 0.0130 | 0.1532 | 0.8783 |
|  | Isotonic | 0.0016 | 0.0141 | 0.2382 | 0.0132 | -0.1841 | 0.8540 |
|  | Sigmoid | 0.0035 | 0.0371 | 0.2181 | 0.0351 | -0.2821 | 0.7779 |

As shown in Figure 6, the Brier Score decreased across most models following post-hoc calibration, confirming improved probability calibration. Random Forest exhibited a marked reduction from 0.007 (uncalibrated) to 0.002 after isotonic calibration. Similarly, XGBoost achieved the lowest overall score (0.002) post-isotonic, despite already having a near-perfect uncalibrated score (0.001), indicating minimal benefit from further calibration. Naive Bayes, initially the poorest performer (0.162), improved substantially to 0.132 after both sigmoid and isotonic adjustments. K-Nearest Neighbors showed a modest but consistent improvement from 0.065 (uncalibrated) to 0.063 under both calibration methods. Logistic Regression and SVM also exhibited slight reductions, with isotonic performing marginally better than sigmoid in both cases. These findings suggest that isotonic calibration offers the most consistent improvement, particularly for tree-based and probabilistic models, though its benefit diminishes when models are already well-calibrated.

**Figure 6.**
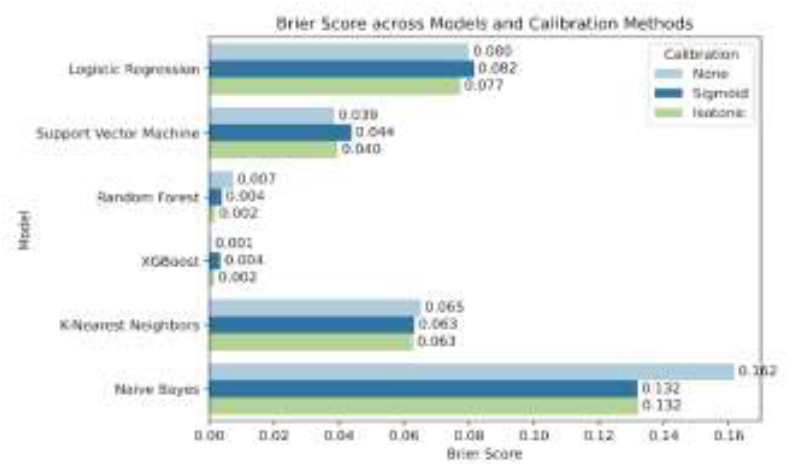
Brier Scores Before and After Calibration

The Expected Calibration Error (ECE) generally decreased after calibration (Figure 7). Notably, Naive Bayes showed the largest reduction, from 0.145 (uncalibrated) to 0.118 (Isotonic). Random Forest similarly improved from 0.051 to 0.011 with isotonic calibration. Support Vector Machine also benefited, reducing from 0.086 to 0.044 post-isotonic. However, Logistic Regression and XGBoost exhibited minimal change, suggesting that they were already well-calibrated. K-Nearest Neighbors (KNN), in contrast, worsened with sigmoid calibration (from 0.035 to 0.081) before slightly recovering with isotonic (0.038). Isotonic regression outperformed sigmoid calibration across most models, demonstrating better reliability in probability estimates.

**Figure 7.**
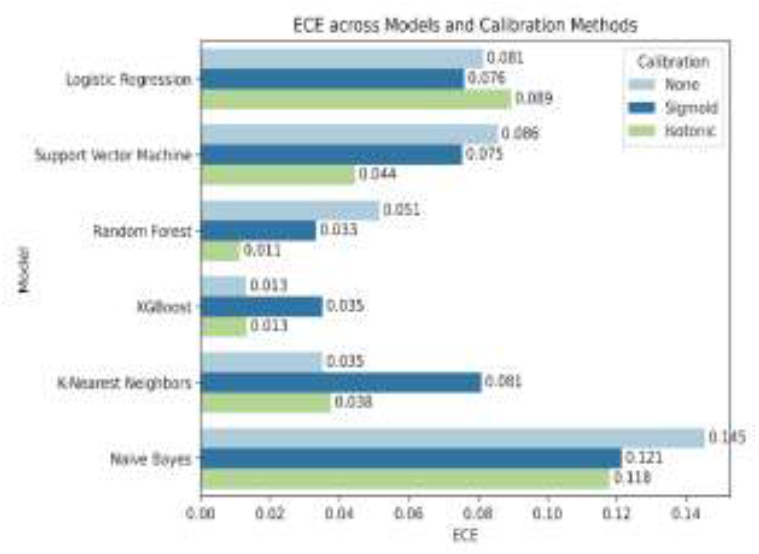
Expected Calibration Error (ECE) Before and After Calibration

Log Loss largely supports the trends observed in Brier Scores (Figure 8). XGBoost exhibited the lowest log loss across all conditions, with minimal variation from 0.013 (None) to 0.014 (Isotonic) reinforcing its strong calibration by default. Random Forest benefited notably from calibration, improving from 0.056 (None) to 0.012 (Isotonic). Conversely, Naive Bayes demonstrated severe miscalibration in its raw form, with an uncalibrated log loss of 1.936, which was substantially reduced to 0.446 following isotonic regression. This highlights its tendency toward overconfident yet incorrect predictions. K-Nearest Neighbors (KNN) also showed calibration sensitivity, where sigmoid calibration worsened performance from 0.183 to 0.207, while isotonic returned it to baseline. Logistic Regression and SVM exhibited modest improvements, with SVM decreasing from 0.142 to 0.133 post-isotonic. Isotonic regression consistently lowered log loss, further confirming its advantage for improving model calibration under most conditions.

**Figure 8.**
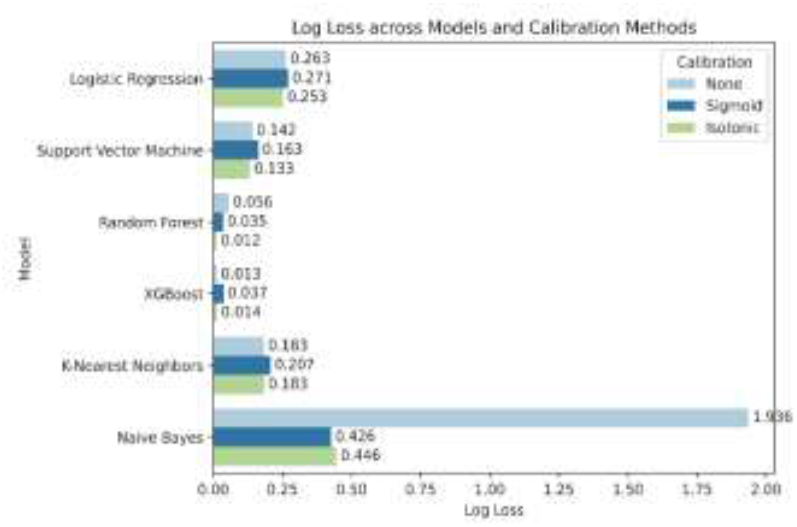
Log Loss Before and After Calibration

Spiegelhalter’s Z-Score and p-value (Figure 9) offer a statistical measure of how well predicted probabilities match actual outcomes [48]. Ideally, a Z-score close to zero paired with a p-value > 0.05 indicates statistically sound calibration. Among all models, Random Forest and XGBoost consistently displayed near-zero Z-scores and high p-values across all calibration settings (p > 0.75), affirming stable and well-calibrated predictions. Naive Bayes, which was severely miscalibrated in earlier metrics, improved substantially after isotonic calibration (Z = 0.30, p = 0.77), compared to its uncalibrated state (Z = -0.60, p = 0.55). Similarly, Logistic Regression achieved near-zero Z-scores (e.g., Z = -0.04 post-isotonic) and strong p-values (≥ 0.88), confirming minimal deviation from expected reliability.

**Figure 9.**
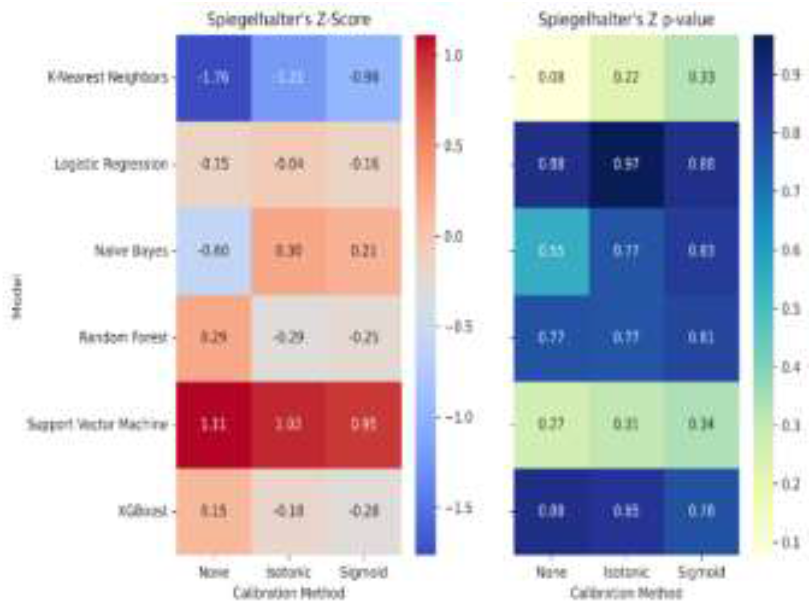
Heatmaps of Spiegelhalter’s Z-score (left) and p-value (right) across models and calibration methods.

K-Nearest Neighbors (KNN) exhibited negative Z-scores across all settings (e.g., Z = -1.21, -0.98), yet its p-values increased with calibration (up to p = 0.33), suggesting partial improvement. Support Vector Machine (SVM) had the highest Z-scores overall (1.02-1.11), and consistently low p-values (≈ 0.27-0.34), implying persistent miscalibration despite adjustments. These results further emphasize that Spiegelhalter’s test provides complementary validation to Brier, ECE, and Log Loss, with isotonic calibration yielding statistically significant reliability gains in most models.

To explore the relationship between calibration and prediction quality, we plotted Expected Calibration Error (ECE) against the Brier Score for all model-calibration combinations (Figure 10). Ideally, well-calibrated and accurate models should lie close to the diagonal line, where ECE and Brier Score are proportionally aligned. This was observed in XGBoost and Random Forest, which consistently clustered near the bottom-left corner, regardless of calibration method, indicating strong predictive reliability and calibration consistency. K-Nearest Neighbors (sigmoid) and Naive Bayes (uncalibrated) appeared well above the diagonal, indicating inflated ECE relative to their Brier Scores. This suggests that while these models may exhibit seemingly reasonable predictive accuracy (as measured by Brier Score), their probability estimates are poorly calibrated, which could undermine trust in probabilistic outputs.

**Figure 10.**
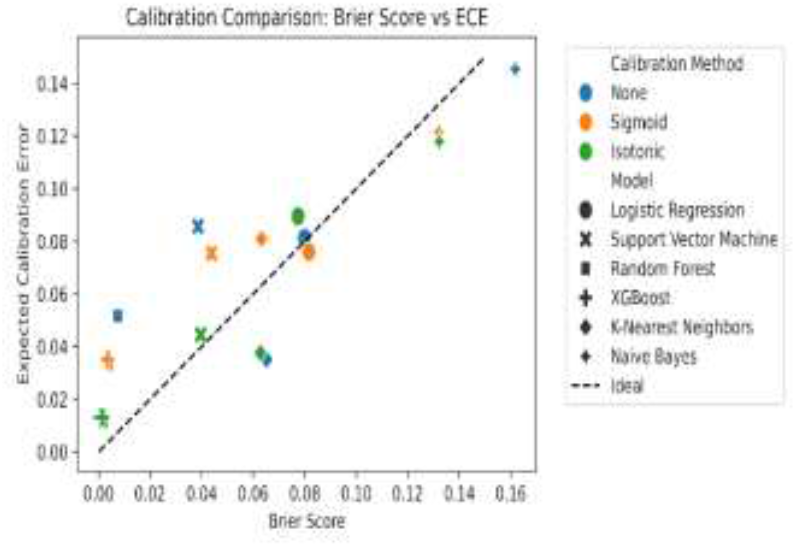
Scatter Plot of Brier Score vs. Expected Calibration Error (ECE) Across All Models and Calibration States

This divergence highlights a key distinction: while Brier Score captures both calibration and classification performance, ECE specifically isolates calibration drift, making it an essential complement in evaluation. These findings support prior recommendations [5] to jointly assess calibration with multiple complementary metrics, thereby providing a multidimensional view of model trustworthiness, especially post-calibration.

### 3.3 Sharpness Analysis

Figure 11 presents the sharpness of model predictions, measured as the variance of predicted probabilities. Higher variance indicates more confident predictions, while lower variance suggests more cautious, uniform output distributions. Random Forest and XGBoost exhibited the sharpest predictions, with Random Forest (Isotonic) reaching the highest variance at 0.240, followed closely by XGBoost (Isotonic) at 0.238 and XGBoost (uncalibrated) at 0.237. These results demonstrate the models’ inherent confidence, which was largely preserved post-calibration. Naive Bayes (uncalibrated) also showed relatively high sharpness at 0.224, but experienced a substantial reduction after calibration, dropping to 0.128 with isotonic and further down to 0.086 with sigmoid. This steep decline in sharpness suggests that Naive Bayes, while initially overconfident, became excessively conservative post-calibration especially under sigmoid, which applied the strongest regularisation.

**Figure 11.**
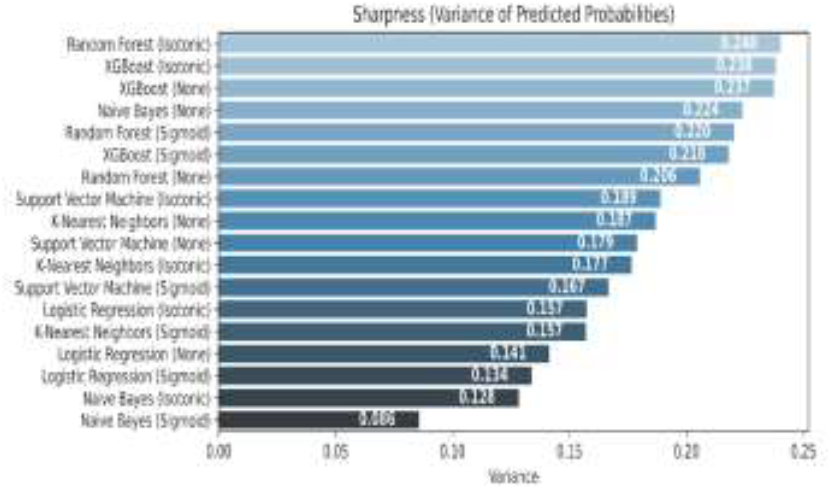
Variance of predicted probabilities (“sharpness”) across models and calibration methods.

Logistic Regression followed a similar trend, with sharpness decreasing from 0.141 (uncalibrated) to 0.134 (Sigmoid) and 0.128 (Isotonic), indicating calibration-induced smoothing. K-Nearest Neighbors and Support Vector Machine exhibited moderate sharpness across all methods, though in both cases sigmoid calibration led to noticeable reductions compared to isotonic. For instance, KNN decreased from 0.187 (uncalibrated) to 0.157 (Sigmoid), and SVM dropped from 0.179 (uncalibrated) to 0.167 (Sigmoid).

## 4 Interpretation of Results

This study demonstrates the significant impact of post-hoc calibration methods on model confidence, calibration quality, and statistical reliability in heart disease prediction. Isotonic regression consistently outperformed sigmoid calibration across most metrics, especially in reducing ECE and Log Loss. For example, Random Forest’s ECE dropped from 0.051 (uncalibrated) to 0.011 (isotonic), with Log Loss reducing from 0.056 to 0.012 (Figure 7-8). Naive Bayes also showed strong gains, with Log Loss improving from 1.936 to 0.446 and its Z p-value increasing from 0.08 to 0.83 (Figure 9), confirming isotonic regression’s effectiveness for overconfident models.

These findings support existing theory that sigmoid calibration, being a parametric method, often suppresses extreme probabilities more aggressively than isotonic calibration, which is non-parametric and locally adaptive. While such smoothing can enhance calibration reliability, it may also trade off against the informativeness of predictions highlighting the need to balance confidence and calibration depending on the deployment context [9].

In clinical applications, where treatment decisions may hinge on predicted risk probabilities, miscalibrated models can yield overconfident but incorrect predictions, potentially leading to under-treatment or overtreatment of patients. For example, Naive Bayes pre-calibration predicted extreme probabilities but had poor alignment with actual outcomes (high Log Loss, low p-value), which post-calibration successfully corrected. This highlights the need for calibration pipelines in AI-assisted diagnostics to improve trustworthiness and reduce the risk of confidence-driven misdiagnosis [56]. Moreover, the use of p-values and Z-scores provides a statistical basis for interpreting model reliability beyond visual tools, such as reliability diagrams.

Previous studies have employed post-hoc calibration in medical machine learning, but many rely solely on a single metric such as Brier Score or ECE [5], [57]. This work expands the scope by combining six (6) complementary metrics: Brier, ECE, Log Loss, Z-score, P-value and Sharpness across six (6) classifiers, offering a more multidimensional view of calibration performance. The incorporation of Sharpness (Figure 11), reflecting the variance of predicted probabilities, adds an important lens into model confidence. For instance, sigmoid calibration consistently reduced sharpness more than isotonic, as observed in Naive Bayes (0.086 post-sigmoid vs. 0.128 post-isotonic), highlighting a trade-off between confidence and calibration smoothness. Few prior studies visualize calibration results this comprehensively across model types. Our comparative heatmaps (Figures 9) and calibration-vs-confidence scatter plots (Figure 10) allow practitioners to assess which models and calibrations offer both reliability and actionable probabilities, a balance rarely presented in calibration literature but essential for healthcare deployment [58].

The contribution of this work lies in demonstrating that calibration effectiveness is highly model-dependent and we present a multi-metric evaluation framework for assessing model calibration in binary heart disease prediction. Our findings show that some models such as Naive Bayes and Random Forest benefit from isotonic calibration, while others such as KNN deteriorate, especially under sigmoid. By introducing sharpness as an evaluative dimension, we go beyond correctness to assess confidence dispersion, an area still underreported in medical AI calibration research. This dual lens of both reliability and confidence is essential for clinical interpretability, risk stratification, and model auditability, especially when deploying decision-support tools in real-world healthcare systems.

This study is limited by the size of the dataset (N=1,025), which may constrain generalizability to broader clinical populations. Although five-fold cross-validation was employed to mitigate variance, the absence of an external validation cohort restricts our ability to assess overfitting or calibration robustness in unseen data. Additionally, we did not test more advanced calibration methods such as Bayesian binning or temperature scaling, which could further refine probability estimates. Future research should explore these techniques on larger datasets and assess calibration dynamics over time or across demographic subgroups for fairness assurance.

## 5. Conclusion

This study evaluated the calibration performance of six classification models for heart disease prediction, using a range of post-hoc techniques and uncertainty metrics. While most models exhibited high accuracy and ROC AUC, their probability estimates often lacked alignment with observed outcomes, underscoring the importance of calibration beyond traditional performance metrics.

Post-hoc calibration particularly isotonic regression demonstrated consistent improvements in reducing Brier Score, Expected Calibration Error (ECE), and Log Loss across models, with Spiegelhalter’s Z-test further validating statistical calibration gains. Compared to sigmoid, isotonic calibration proved more effective for correcting non-linear miscalibrations, especially in probabilistically unstable models such as Naive Bayes and ensemble classifiers. However, models such as KNN remained challenging to calibrate regardless of method, highlighting the limits of post-hoc approaches for certain decision boundary behaviors.

These findings reinforce the need to assess and calibrate predictive uncertainty when deploying machine learning models in clinical settings. Well-calibrated probabilities are essential for accuracy, for supporting trustworthy decision-making and risk communication. Given its flexibility and empirical advantage, isotonic regression is recommended as the preferred calibration method in healthcare AI applications.

## Data Availability

All data produced in the present work are contained in the manuscript

## Data and Code Availability

The datasets analyzed during the current study is publicly available at Kaggle (https://www.kaggle.com/datasets/johnsmith88/heart-disease-dataset). The code supporting the findings of this study is available from the corresponding author on reasonable request.

## About the authors

Peter Adebayo Odesola holds a master’s degree in Artificial Intelligence and Data Science from Solent University (2022) and a postgraduate diploma in Education. His expertise spans data analytics, machine learning, and AI in healthcare, with projects on predictive modelling, automation, and visualisation that advance data-driven solutions in both academic and professional contexts.

Adewale Alex Adegoke holds a master’s degree in Applied Artificial Intelligence and Data Science from Southampton Solent University, completed in 2023. During his time at the university, he contributed to various innovative research initiatives. Adewale has since worked as a Data Scientist at 10Alytics, where he applied advanced technologies and research methodologies to craft data-driven solutions for businesses. He currently serves as a Data Systems Manager at the Westminster Foundation for Democracy (WFD), UK. His research focuses on the application of advanced data analytics and machine learning techniques to solve real-world challenges.

Idris Babalola is a Senior Data Scientist with the Department of Health and Social Care, UK. He has held previous part-time roles at Solent University United Kingdom as a Data scientist, MSc research supervisor as well as Associate lecturer in Computing. He holds a MSc in AI and Data science from Solent University (2022). His research interest lies in the use of AI for Healthcare utilising data science skills, NLP and large language models.

